# Detecting and understanding meaningful cancerous mutations based on computational models of mRNA splicing

**DOI:** 10.1101/2023.12.05.23299582

**Authors:** Nicolas Lynn, Tamir Tuller

## Abstract

Cancer research has long relied on non-silent mutations. Yet, it has become overwhelmingly clear that silent mutations can affect gene expression and cancer cell fitness. One fundamental mechanism that apparently silent mutations can severely disrupt is alternative splicing. Here we introduce *Oncosplice*, a tool that scores mutations based on models of proteomes generated using aberrant splicing predictions. *Oncosplice* leverages a highly accurate neural network that predicts splice sites within arbitrary mRNA sequences, a greedy transcript constructor that considers alternate arrangements of splicing blueprints, and an algorithm that grades the functional divergence between proteins based on evolutionary conservation. By applying this tool to 12M somatic mutations we identify 8K deleterious variants that are significantly depleted within the healthy population; we demonstrate the tool’s ability to identify clinically validated pathogenic variants with a positive predictive value of 94%; we show strong enrichment of predicted deleterious mutations across pan-cancer drivers. We also achieve improved patient survival estimation using a proposed set of novel cancer-involved genes. Ultimately, this pipeline enables accelerated insight-gathering of sequence-specific consequences for a class of understudied mutations and provides an efficient way of filtering through massive variant datasets – functionalities with immediate experimental and clinical applications.

## INTRODUCTION

Advancements in sequencing technology have made extensive collections of mutations and genomic information available^1–4^. These datasets include millions of novel mutations that cannot all be experimentally studied due to time and cost constraints. Thus, most investigations that characterize functional variants focus on non-silent, non-synonymous mutations^5,6^. This heuristic effectively narrows the search space yet neglects thousands of apparently silent mutations with measurable and potentially more severe consequences. Instead of directly altering coding nucleotides, silent and apparently silent mutations act on regulatory gene expression processes^6–10^; they can exist within introns and untranslated regions, or within coding sequences (CDS)^3–11^, and they hold significant predictive power in cancer classification and prognosis^11^. Among the regulatory mechanisms that they can hijack is splicing^9,12–25^.

mRNA splicing is a co-transcriptional modification step that transforms one pre-mRNA sequence into multiple transcripts through alternative splicing (AS). The most crucial *cis-*elements needed for proper splicing are the intron’s 5′ (donor – GU motif) and 3′ (acceptor – AG motif) junctions. There are also hundreds if not thousands of sequence determinants far within and beyond the intron that are more difficult to characterize and play roles of varying importance in deciding which GU/AG dinucleotides serve as functioning splice sites^26^. Ultimately, this gives cancerous apparently silent mutations countless targets through which they can disrupt healthy gene expression.

Numerous examples illustrate the impact of aberrant splicing in cancer^15,27–29^. One estimate claims that between 15% and 50% of human disease mutations can result in splicing dysregulation^24^. It was found that 68% of tumor samples contained at least one aberrant splicing-derived neoepitope while only 30% contained neoepitopes derived from somatic single-nucleotide variants^30^. Exons 4, 6, and 9 of *TP53* contain functional hotspots for intron retention-caused inactivation by SNVs, and mutations causing such effects are visible in lung squamous cell carcinoma^31^. The Warburg effect exercised by some tumors depends upon a shift in expression from adult-observed pyruvate kinase isoforms to embryonic-observed splicing patterns^15,31–33^. Even tumor drug resistance is linked to splicing, as shown with a *vemurafenib*-resistant isoform of *BRAF* that is lacking exons 4-8^23,34^.

Due to the clear relevance of splicing in cancer and other disease phenotypes, several pathogenicity predictors related to splicing have been published. These include tools such as *CADD*^35^*, MMSplice*^36^*, TraP*^37^*, IntSplice2*^38^*, RegSNPs-Intron*^39^*, RegSNPs-Splicing*^40^*, and S-CAP*^41^; these models typically employ machine learning (ML), use training procedures to classify deleterious mutations based on *a priori* knowledge of pathogenicity, provide limited mechanistic insight, and are constrained to specific mutation types and regions. These tools can help identify likely pathogenic mutations but offer limited information related to the functional characteristics and splicing events that make their detected mutations deleterious.

To computationally elucidate mutations’ effects through missplicing, we propose *Oncosplice*. This pipeline predicts aberrant splicing events, generates variant transcriptome annotations, constructs resultant proteomes, and provides a measure of pathogenicity based on estimated functional divergence measurements between reference and aberrant proteins. We demonstrate that the tool captures multiple meaningful signals in a large set of somatic, unannotated mutations obtained from The Cancer Genome Atlas (TCGA) Program, can outperform other splicing-related pathogenicity predictors on *ClinVar* variants, and improves patient survival estimation.

## RESULTS

### 1. Approximately 1.3% of all somatic mutations in cancer patients may cause aberrant splicing

We analyzed 12,250,236 unique somatic mutations across 9,874 genes from 8,362 patients with any of 19 cancer types accessed from and curated by TCGA. We found that 159,458 variants result in at least one predicted missplicing event. Specifically, these mutations induced 51,066 missed acceptors, 43,924 missed donors, 40,844 discovered acceptors, and 26,463 unique discovered donors. 64,368 mutations resulted in two or more missplicing events. Moreover, 8,179 missplicing mutations receive *Oncosplice* scores ≥ 2,000 (top 5^th^ percentile), a threshold representing variants with especially deleterious changes to the proteome. We refer to these variants as predicted deleterious mutations. We estimate that at least 1.3% of somatic variants in tumors result in some missplicing.

The most easily identifiable missplicing mutations are those that alter core splicing motifs, and, as can be seen in **Figure 2A**, splice site mutations account for 62,047 or 38.9% of all predicted missplicing mutations. Meanwhile, of all 65,171 splice site mutations analyzed, 3,124 were not detected to cause missplicing. They are likely missed since many splice sites are used alternatively; as non-constitutive junction usage is correlated with lower *SpliceAI* probabilities^26^, the deletion of such splice sites would be characterized by changes in *SpliceAI* probability that are smaller than the detection threshold used. We can see where splice site-deleting deleterious SNVs reside relative to their deleted junctions in **Figure 2F**, and among acceptor-deleting mutations, 55% exist at the splice site they delete. However, among donor-deleting variants, only 33% reside at the splice site they delete, 26.8% are missense mutations one nucleotide into the exon from the deleted donor, and 22.7% reside more than 7 nucleotides upstream of the site; while selection of splice site mutations as deleterious is significant (p value: 0.018, 0.006 for acceptors and donors respectively), there are several interesting mutations outside the focus of splice junctions. Therefore, understanding splice site mutations alone does not adequately represent the space of meaningful missplicing mutations.

**Figure 1.**
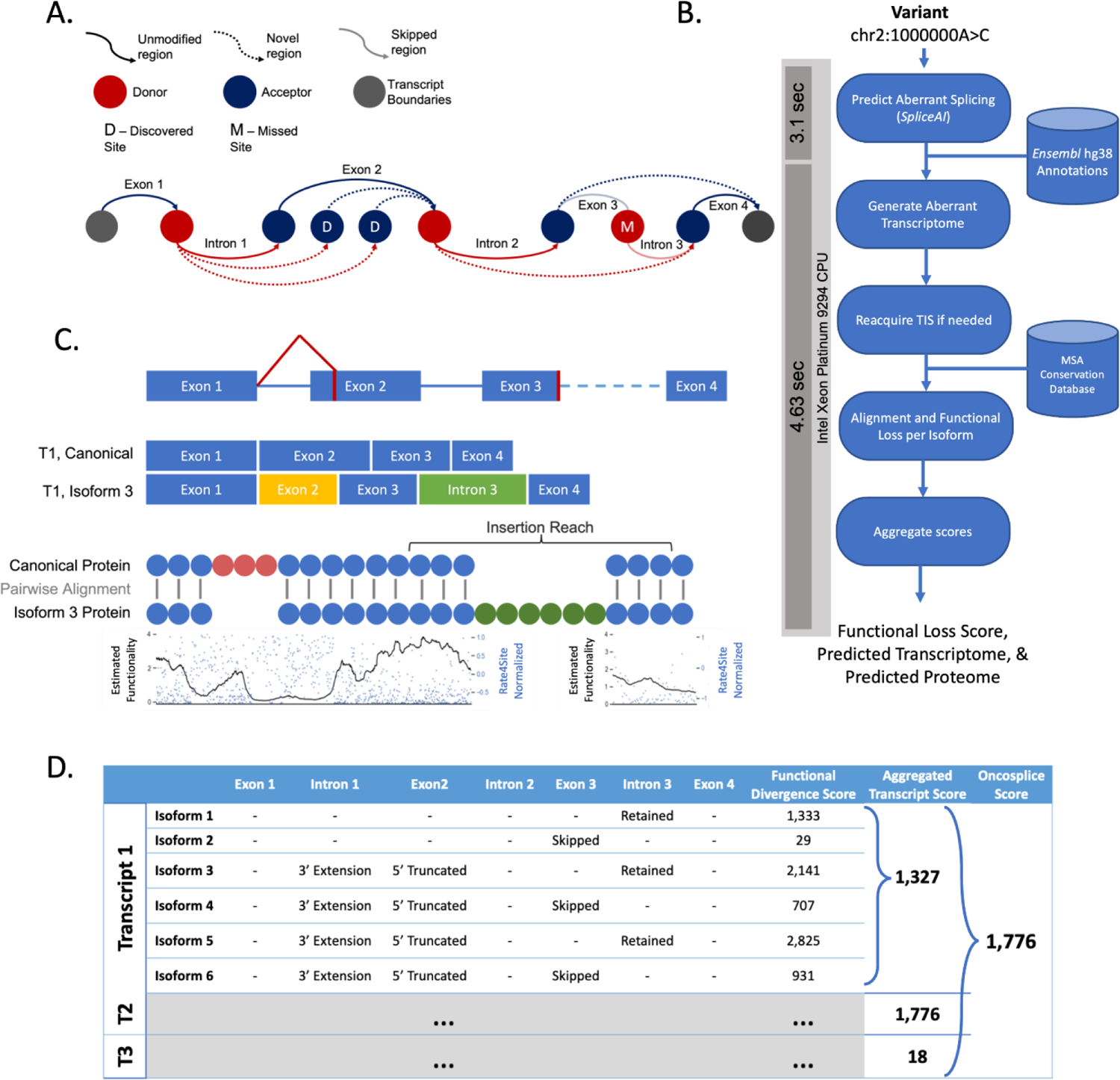
Description of the *Oncosplice* pipeline. A. A diagram illustrating the greedy approach to constructing transcript isoforms given only a pool of splice sites with predicted missplicing such as missed or cryptic junctions. B. Overview of the steps taken in the pipeline to obtain a concise quantitative description of the functional loss that a mutation induces through predicted missplicing, and the transcriptome annotations; general run-time per mutation consumes 7.73 seconds when running on an Intel Xeon Platinum 9294 CPU, with 40% of the time spent running SpliceAI inference and 60% spent predicting, generating, and grading variant transcriptomes. C. Comparing two proteins using conservation scores per amino acid using an algorithm that captures the loss due to insertions and deletions. D. Mature mRNA sequences are translated by selecting TISs with more optimal context based on TITER, Kozak context, and folding in the case that TISs are interrupted by mutations or by splicing. E. A view of the collected data for each transcript’s isoforms, and the aggregation of functional loss scores at the transcript and gene level.

**Figure 2.**
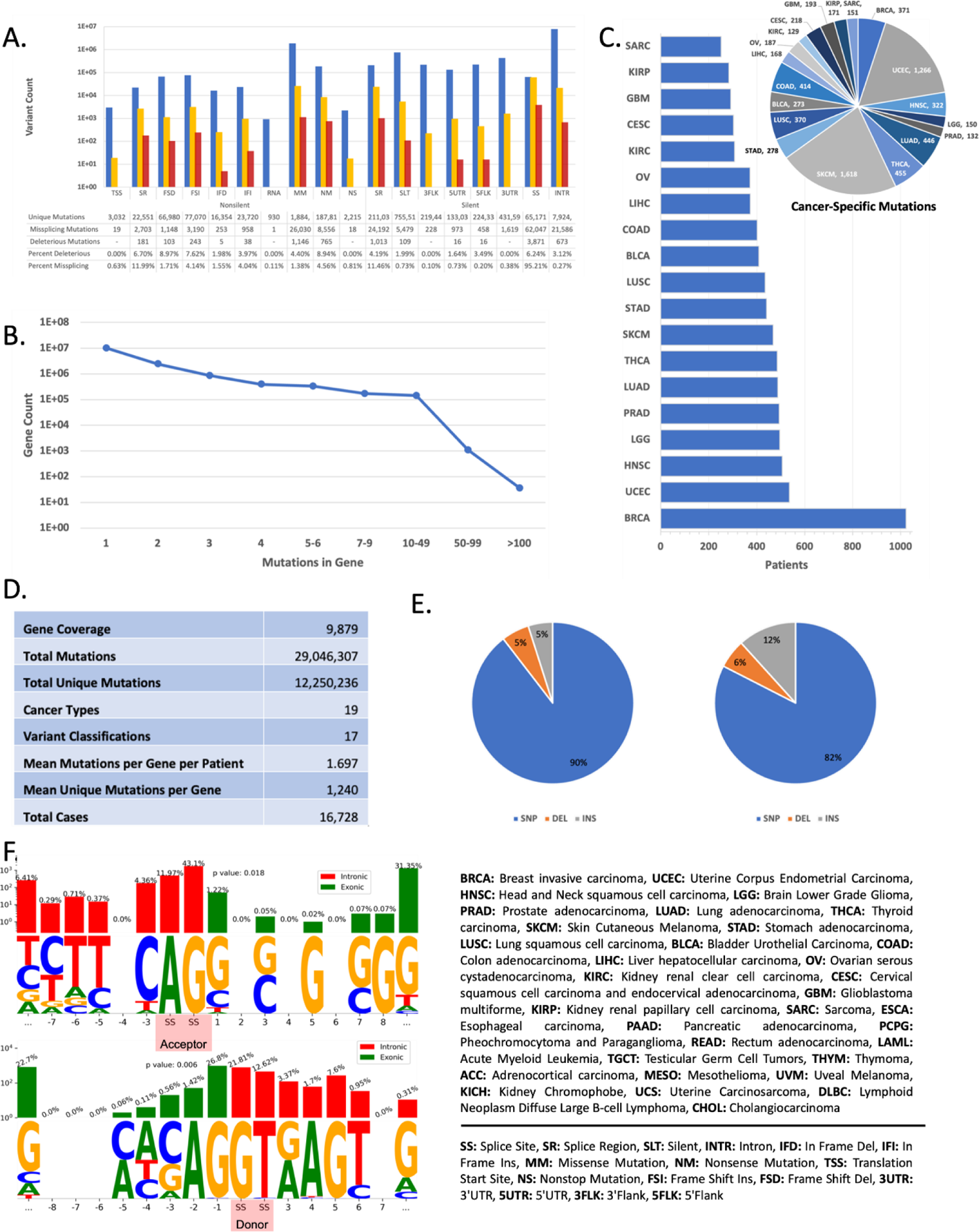
Reference dataset statistics. A. The proportion of all mutations per variant classification that are retained in the missplicing and deleterious missplicing subsets reveals that predicted deleterious mutations come from all regions, and more significantly from splice sites. B. A plot describing the distribution of variants per gene per patient shows that typically genes have between 1 and 2 mutations. C. A breakdown of the cancer types analyzed and how many patients each project includes, with BRCA being the largest in terms of cohort size; the number of cancer-specific deleterious mutations in each cancer type is also displayed (cancer-specific mutations are variants found only among patients with one cancer type). D. General dataset statistics across multiple variant descriptors show that the data processed with Oncosplice is highly diverse. E. The proportion of all unique mutations in each variant type category in the TCGA set available and in the predicted deleterious subset indicates that most somatic mutations analyzed are SNVs, while insertions seem to proportionally induce more splice site alterations as is indicated by their higher composition among deleterious variants. F. Most of the deleterious mutations that induce a missed acceptor fall on or around the splice site motif, as do most of the mutations that induce a missed donor, though there are several variants that disrupt both junctions from hundreds of nucleotides away.

Splice region mutations (within 3-8 bases of the intron or within 1-3 bases of the exon) and non-silent coding variants also account for 67,068 missplicing variants. Of those, 36,030 are missense mutations (16% of predicted missplicing mutations), as seen in **Figure 2A**, demonstrating how the most widely examined class of mutations may have secondary consequences related to splicing beyond their conspicuous and distracting amino acid exchanges. 8,556 nonsense mutations (5% of missplicing mutations), which generate early termination codons, also result in missplicing which possibly neutralizes their otherwise truncating effects via partial or full exon skipping and intron retention. For example, one identified insertion mutation induces an early stop codon but also contains a cryptic donor site within the context of the inserted sequence. The cryptic donor relegates the stop codon to a novel intronic region and preserves the rest of the protein while only deleting a segment of 2 amino acids. This mutation affects six patients with thyroid cancer and is not seen in the general population.

Interestingly, 8,417 intronic mutations were found to delete at least one splice site, while 16,776 intronic mutations were found to generate at least one cryptic splice site. Detection fidelity decays for deeper intronic variants when using whole exome sequencing (WES), and this class is likely underrepresented in this study. Still, intronic variants accounted for 13.54% of missplicing variants and 673 (8.2%) of deleterious variants, further highlighting the functional value of understanding intronic mutations in disease. Finally, silent exonic variants, which are very easily overlooked, accounted for 5,479 missplicing mutations and 109 predicted deleterious variants.

### 2. Predicted misspliced transcripts agree with isoforms causally linked to mutations in RNAseq-based studies

Several computational investigations have estimated the causal relation between somatic SNVs and missplicing events such as cryptic splice junction creation, exon skipping, and intron retention. This can be done by isolating genes with single mutations and detecting aberrantly spliced reads via RNAseq. In these investigations, thousands of mutations were connected to specific splicing outcomes using allele-specific or ratio-based splicing analysis^31,42^ or junction allele fractions (JAF)^43^.

First, we processed 1,152 mutations that were reported by *MiSplice*^43^ to induce novel intronic splice junctions. After lifting the novel splice sites to the *hg38* genome build, we checked these cryptic splice junctions to those predicted computationally with *Oncosplice*. Out of 1,152 variants, 109 mutations were not predicted to cause missplicing. The remaining 1,043 (90.54%) mutations were found to cause a cryptic splice site at a *SpliceAI* threshold of 0.25, and the *MiSplice-*identified position was always less than 3 nucleotides from the computationally predicted site. Of those, 723 variants caused a single cryptic acceptor while 313 caused a single cryptic donor, and 7 variants caused both a cryptic donor and acceptor together. Furthermore, each mutation had JAF describing the proportion of reads mapped to a relevant transcript region that expressed the cryptic splice junction. These splice junction JAFs significantly correlated with computationally predicted probabilities, as can be seen in **Figure 3A-B**. This finding reinforces the utility of *SpliceAI* probabilities as proxies for splice site penetrance.

**Figure 3.**
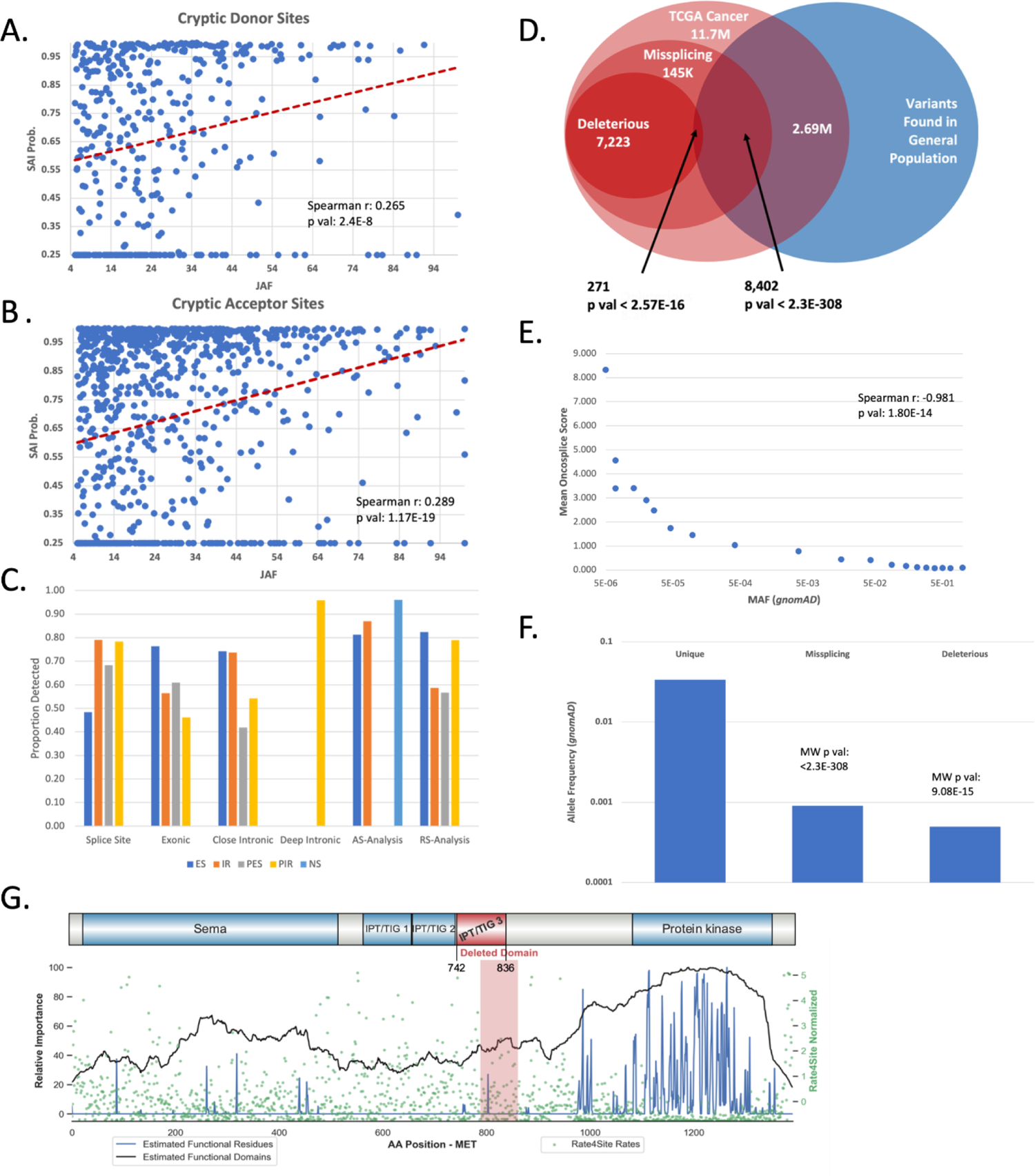
Benchmarking *Oncosplice* predictions using RNAseq and allele frequencies. A-B. Plotting the junction allele fraction for cryptic splice sites induced by variants against their *SpliceAI* probability reveals a significant correlation between the latter and discovered splice junction penetrance (as measured using RNAseq and *MiSplice*); we used a *SpliceAI* detection threshold of 0.25 and any site with a change in penetrance below this is not detected. Moreover, we only observed the positions for which the lifted hg38 coordinate of the *MiSplice*-identified cryptic splice site was within 3 nucleotides of the *SpliceAI*-identified cryptic splice site. C. We calculate the discovery ratio as the proportion of splicing events identified in two separate RNAseq-based computational investigations that were properly predicted by *Oncosplice.* D. A depletion of variants occurring in the general population among predicted deleterious mutations indicates the added insight *Oncosplice* generates on top of simply identifying missplicing mutations with *SpliceAI*. E. We analyzed the mean *Oncosplice* score of variants binned based on *gnomAD* MAF into similarly sized sets (∼1.4E4 mutations per bin) and reveal a significant correlation. F. The mean *gnomAD* MAF is significantly lower among predicted deleterious variants than among missplicing variants, and significantly lower among missplicing variants than among non-missplicing variants. G. A splice site mutation in MET’s 10^th^ intron results in a skipped exon and deletes a large part of the protein’s functional domain.

Next, we compared the results of allele-specific splicing analysis on *RNAseq* data – which identified events including exon skipping (ES), intron retention (IR), partial exon skipping (PES), and partial intron retention (PIR) – to the transcript isoforms estimated and constructed by *Oncosplice*. Specifically, we analyzed 761 mutations identified through ratio-based splicing analysis^31^, 219 mutations identified through allele-specific association^31^, and 267 close intronic, 459 splice site, 286 exonic, and 228 deep intronic variants identified using a combination of read-ratios and allele specificity^42^. Of these mutations processed, we retained for analysis those that were predicted to induce a missplicing event at a detection threshold of 0.25. Since *Oncosplice* predicts multiple isoforms, for each variant we recorded whether *Oncosplice* predicted the event detected in RNAseq. As summarized in **Figure 3C**, in 11 of 20 categories, *Oncosplice* identified more than 70% of the isoforms observed. Events induced by splice site mutations are most effectively reconstructed, in large part due to the increased prediction fidelity that *SpliceAI* provides closer to splice junctions. This is significant, considering *Oncosplice* produces these estimates fully computationally. Splice site usage and *RNAseq* data are highly dependent on factors such as source tissue type, making it likely that many other isoforms predicted with *Oncosplice* exist and were simply not visible in these *RNAseq* experiments.

To understand the events detected by *Oncosplice* more tangibly, we explore two relevant and documented case studies. First, we consider MET, which encodes a receptor protein kinase and is a well-known cancer driver. MET is represented in the TCGA dataset by 2,062 unique mutations. Of those, 34 mutations (1.65%) are predicted to result in missplicing, while only one is scored as pathogenic by *Oncosplice:* chr7:116739948A>T. This variant is predicted to destroy the canonical acceptor of the 10^th^ intron and partially destroy the canonical donor of the 11^th^ intron in MET’s primary transcript. *Oncosplice’s* predicted outcome of this event is exon 11 skipping, which otherwise preserves the protein but deletes 73 amino acids that map to a *TIG* functional domain. Interestingly, this mutation affects 11 patients, 10 of whom belong to the GBM (glioblastoma multiforme) cohort. This variant is classified as likely benign in *ClinVar* and COSMIC, and is associated with renal cell carcinoma, but we found no experimental data or functional evidence describing this mutation’s effects, indicating it has likely not yet been studied. Another set of 14 unique mutations in the vicinity of MET’s 14^th^ intron (a known hotspot for exon skipping variants) is predicted to delete the nearby donor site and ultimately result in full or partial exon skipping leading to a loss-of-function event. These mutations did not meet the established *Oncosplice* threshold for pathogenicity. Out of 15 patients affected by these mutations, 10 belong to the LUAD (lung adenocarcinoma) cohort, a pathology that is well-linked to MET exon 14 skipping^44^. We also processed a KRAS variant known to be involved in *Osimertinib*-resistant lung cancer: chr12:25227343G>T. This mutation causes an amino acid substitution with gain-of-function effects through activation of ERK1/2 activation and RAS-GTP^44^. Yet, this same mutation was experimentally found to induce a cryptic splice site which causes a frameshift and protein truncation wherein the c terminus of the resulting proteins was minimally detectable using antibodies^44^. This event is captured quickly and effectively by *Oncosplice*. The cryptic splice donor is highly penetrant (*SpliceAI* Delta: +0.744) and the protein truncating event is constructed and predicted as the primary isoform with a score of 1,154.

Similarly, *Oncosplice* detected 33 missplicing mutations out of 202 unique variants surrounding exon 4 of TP53 – another known hotspot – that resulted in partial or full exon skipping.

### 3. Predicted deleterious missplicing mutations are significantly depleted within the general population

We used VEP to annotate all SNVs and deletions with combined population allele frequency (AF) data from the *1000 Genome Project*^3^ and *gnomAD*^45^. Frequencies associated with these TCGA mutations vary significantly; some are *de novo* (they are not previously studied or seen in either of these auxiliary sets), while 2.49% of somatic SNVs are observed as the primary allele in the general population (somatic variants are alleles that differ in tumor samples relative to healthy cells from a single cancer patient so, while rare, a patient’s alternate allele can be predominant in the general population). We expect that some mutations found within the general population can be deleterious^46^ and that a vast proportion of variants found in cancer patients are passengers rather than drivers, though typically deleterious and benign variants have lower and higher AFs, respectively. Among all somatic mutations studied, 2.69M variants are found at non-zero AFs in *gnomAD*. These variants are diverse across all descriptors.

First, we tested the depletion of variants occurring in the healthy population within the missplicing and predicted deleterious subsets of variants, a concept illustrated in **Figure 3D**. Of the 144,652 missplicing variants with AFs, only 8,402 are seen in the healthy population (permutation test mean: 33,440, permutation p-value: < 0.001, hypergeometric p-value: < 2.3E-308, Chi-square p-value: < 2.3E-308) indicating that missplicing mutations are more frequent in pathologies than in the healthy population. Of the 7,223 deleterious missplicing mutations with AFs, only 271 are seen in the general population (permutation test mean: 420, permutation p-value: < 0.001, hypergeometric p-value: 2.57E-16, Chi-square p-value: 6.37E-14), a strong depletion calculated against the missplicing mutation set which implies that *Oncosplice* scores contribute significant additional information past predicting the occurrence of aberrant splicing. Missplicing variants not seen in the general population receive more pathogenic scores than healthy-observed missplicing mutations (difference: 132, permutation random mean: −0.002, p-value: < .0001, Wilcoxon Rank Sum: 8.66E-83). As seen in **Figure 3E**, there is also a significant negative correlation between *gnomAD* AFs and mean *Oncosplice* scores. Since we expect that missplicing mutations in the healthy population would generally have less severe disease-related effects, this further suggests that *Oncosplice* scores accurately convey the nature of a variant’s functional consequences.

### 4. *Oncosplice* outperforms alternative splicing-related pathogenicity predictors and provides actionable insights

Our next aims are to measure the predictive value that *Oncosplice*’s functional divergence algorithm provides in terms of capturing deleterious changes to proteins, benchmark *Oncosplice* scores using other pathogenicity predictors, and establish a threshold that can be used to define deleterious mutations from *Oncosplice* scores. To this end, we download 1,475,305 mutations from the *ClinVar* dataset to analyze the significance of *Oncosplice*’s pathogenicity predictions against clinically validated pathogenicity associations^47–49^. Of those, 398,489 mutations overlapped our processed TCGA variants. We graded all these variants using *Oncosplice’s* functional divergence algorithm, including those that do not result in predicted missplicing. As can be seen in **Figure 4B**, 8.81% of these variants are labeled as pathogenic or likely pathogenic while approximately 48.5% are benign or likely benign. The rest have conflicting interpretations of pathogenicity or lack sufficient clinical evidence to assign a label. 10,833 *ClinVar* mutations result in predicted missplicing. 54.0% of this pool has evidence of pathogenicity while 6.64% are benign (permutation p-value: < 0.001); predicted missplicing variants obtain a positive predictive value (PPV) for pathogenicity of 89%.

**Figure 4.**
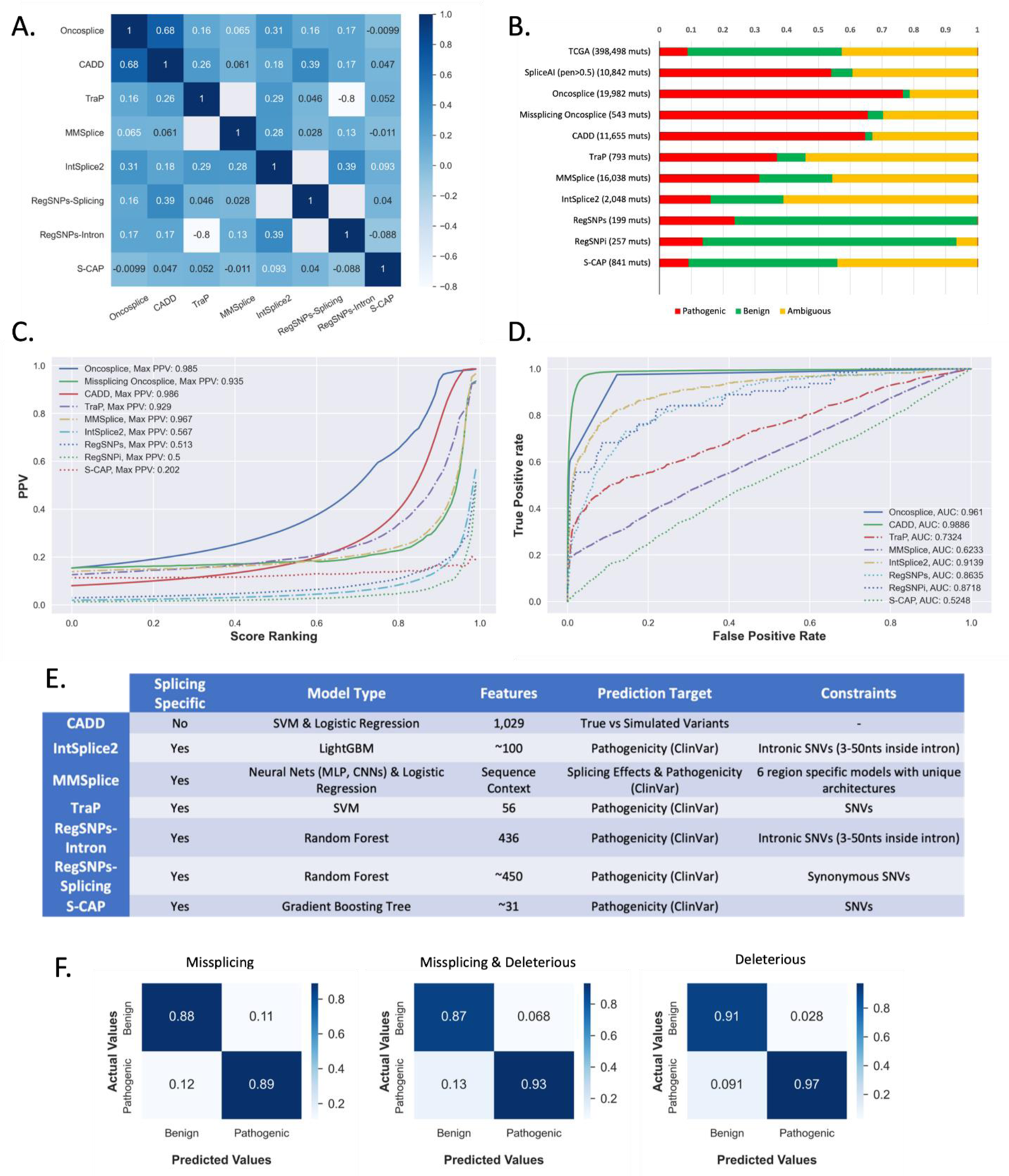
Comparing pathogenicity predictor performances in splicing. A. A heatmap of each tool’s Spearman correlation to all other predictors; tools such as CADD exhibit high correlation to several tools – especially *Oncosplice –* while others such as S-CAP and *Oncosplice* are less correlated indicating they may capture orthogonal information. B. Ratio of pathogenic, benign, and ambiguous variants found in *ClinVar* for subsets of top 5% of predicted deleterious mutations using eight pathogenicity predictors’ scores. C. The positive predictive values for incremental percentiles for each of 8 tools’ scores indicate that *Oncosplice* functional divergence can generate meaningful and narrowed search spaces on parr to *TraP* and *MMSplice*. D. ROC analysis of all the tools indicates that on the task of pathogenicity performance, the scoring algorithm employed by *Oncosplice* is effective, with *CADD* being the only model to outperform in terms of AUC. E. A tabular description of each alternative pathogenicity predictor, along with any notable constraints and training scheme employed. F. Confusion matrices for showing the predictions using binary thresholds based on *Oncosplice* scores for all mutations and missplicing mutations, as well as the prediction quality for considering missplicing mutations as pathogenic alone.

Many other splicing-related pathogenicity predictors have been published. These tools typically leverage machine learning strategies, train directly on pathogenicity classifications (often from *ClinVar,* information *Oncosplice* infers without training) to predict consequences based on *a priori* knowledge of pathogenicity, provide limited to no mechanistic insight, and are often constrained to specific mutation types (synonymous SNVs, missense SNVs) and regions (intronic, splice site, splice region)^35–41^. A tabular description of these tools is available in **Figure 4E**. We compare the results from *Oncosplice* as an end-to-end pathogenicity predictor to results obtained from *RegSNPs-Splicing, Reg-SNPs-Intron, S-CAP, TraP, MMSplice,* and *IntSplice2*. We also compare against *CADD* even though it is not a splicing-specific model that uses hundreds of other features relating to motifs, conservation estimates, and evolutionary mechanisms; it also uses features generated with *SpliceAI* and *MMSplice*. We scored or obtained pre-computed sets of mutations from all alternate models. We find *CADD* and *Oncosplice* scores share the greatest correlation, yet several models are not highly correlated to each other, as shown in **Figure 4A**, indicating some orthogonality in splicing-related predictions.

First, we calculated PPVs for each tool at incremented percentiles to understand prediction quality at different scores. As seen in **Figure 4C**, only *MMSplice*, *TraP*, and *CADD* reach levels as high as *Oncosplice.* We show results for *Oncosplice* scores when masking out variants that aren’t predicted to cause missplicing (isolating predictions for missplicing variants) as well as for all mutations together (to understand the scoring algorithm’s capture of functional changes). It is interesting to note that the functional divergence score outperforms most tools and even provides a larger search space with an equally high PPV as *CADD* at weaker percentiles. For a clear view of these predictions, we show the clinical significance labels associated with the variants in the top-scoring 95^th^ percentiles for each model. As shown in **Figure 4B**, no other predictor identifies pathogenic mutations at ratios as high as with *Oncosplice* both for missplicing mutations and all mutations. We also compare the performance of all tools using receiver operating characteristic (ROC) curves, despite their being a less suitable metric for the task of splicing-related pathogenicity identification due to the expectedly large quantity of falsely labeled benign mutations (which are simply mutations that are pathogenic through mechanisms unrelated to splicing); this is done to test the significance of *Oncosplice’s* scoring algorithm and its independence of splicing. It is a fair measure as all other predictors used supervised machine learning algorithms to directly predict *ClinVar* classifications and, therefore, have seen the data we measure performance against. In **Figure 4D**, the ROC performance of *Oncosplice*’s protein comparison algorithm approaches that of *CADD*, a state-of-the-art tool in pathogenicity prediction. Notably, *Oncosplice* and *CADD* can achieve top predictive performance without training on pathogenicity at all. Because we do not use a training scheme in constructing *Oncosplice*, we can also guarantee that its performance is not affected by data circularity that may affect ML-leveraging models^38^.

Additionally, *Oncosplice* provides insight into missplicing mutations that are ORF-bound and non-synonymous, which no other model handles. These mutations may have distracting and direct effects on the amino acid composition but may have secondary effects on splicing.

Similarly, recent investigations point to UTR variants’ role in missplicing. Several of the mutations *Oncosplice* identifies as deleterious reside in the 5’UTR region, and because *Oncosplice* provides estimated transcriptomic and proteomic libraries we can use this tool to study their effects further. Pathogenicity prediction is well established, and most methods offer high prediction performance, yet methodology now becomes increasingly important as we try to computationally understand the mechanisms through which variants are harmful; *Oncosplice* is the only model considered that offers so much in this regard. Ultimately, we show *Oncosplice* performs competitively in the task of pathogenicity prediction without the central reliance on ML as a score generator, without prior knowledge of pathogenicity, without the need for a training or optimization scheme, and without variant constraints, all as a secondary task to proteome estimation.

When looking at the set of variants that meet an *Oncosplice* threshold of 690, or in the top 95^th^ percentile for missplicing variants, we find 542 missplicing variants, 65.5% of which are pathogenic and 4.5% of which are benign (PPV value: 93.17%, permutation p-value: < 0.001). Therefore, we establish the top 5% of missplicing variants by *Oncosplice* score among arbitrary sets of analyzed mutations as those most likely to be pathogenic. Among the phenotypes associated with the pathogenic mutations identified with *Oncosplice* are several cancer-related terms, including hereditary cancer predisposition syndrome, familial cancer of breasts, breast-ovarian cancer, gastric cancer, and colorectal cancer.

### 5. Genes overrepresented with deleterious mutations are enriched with known cancer drivers and reveal novel biomarkers that improve patient survival estimates

There are several published lists of classical cancer drivers^50–58^. These lists are often based on non-silent mutations, can be developed either through computational or experimental investigations, and ultimately enable targeting for treatment development. If *Oncosplice* functions properly, it can be reasoned that many of those genes overrepresented with deleterious missplicing mutation are known cancer drivers due to direct selection within a cancer cohort. To this end, we search for deleterious mutation-overrepresented genes using hypergeometric probability.

To identify significant genes while controlling for selection bias related to total mutation volume, we group genes into 5 distinct bins within each of which selected genes and background genes have insignificantly different mutation volumes, and then rank genes in each bin by the ratio of deleterious missplicing mutations to all unique mutations. We then scan through the top percentiles across all bins and assess the identification of drivers. More details on this procedure are available in the Methods. As can be seen in **Figure 5A**, there is a strong enrichment of pan-cancer driver genes, which reportedly play underlying roles in multiple pathologies. We also test for the enrichment of known TSGs and oncogenes separately using the same procedure and role-specific gene sets. It is seen in **Figure 5A** that TSGs are enriched more strongly than oncogenes, indicating either that missplicing is a more typical precursor in TSG inactivation than in oncogene gain-of-function, or that the scoring strategy implemented better captures behaviors typical of TSG knockout. Quantifying novel protein functionalities that cause an upregulation of activity or change of functionality is a much more difficult task. We also perform enrichment of pan-cancer drivers in sets of genes that are overrepresented in cancer-specific variant subsets. Moreover, we compare the enrichment of these identified drivers against drivers identified while checking for overrepresentation in the missplicing subset. As can be seen in **Figure 5B**, cancer drivers are much more strongly enriched among genes overrepresented by deleterious mutations compared to genes overrepresented by missplicing mutations, reinforcing the added value of *Oncosplice* on top of *SpliceAI*.

**Figure 5.**
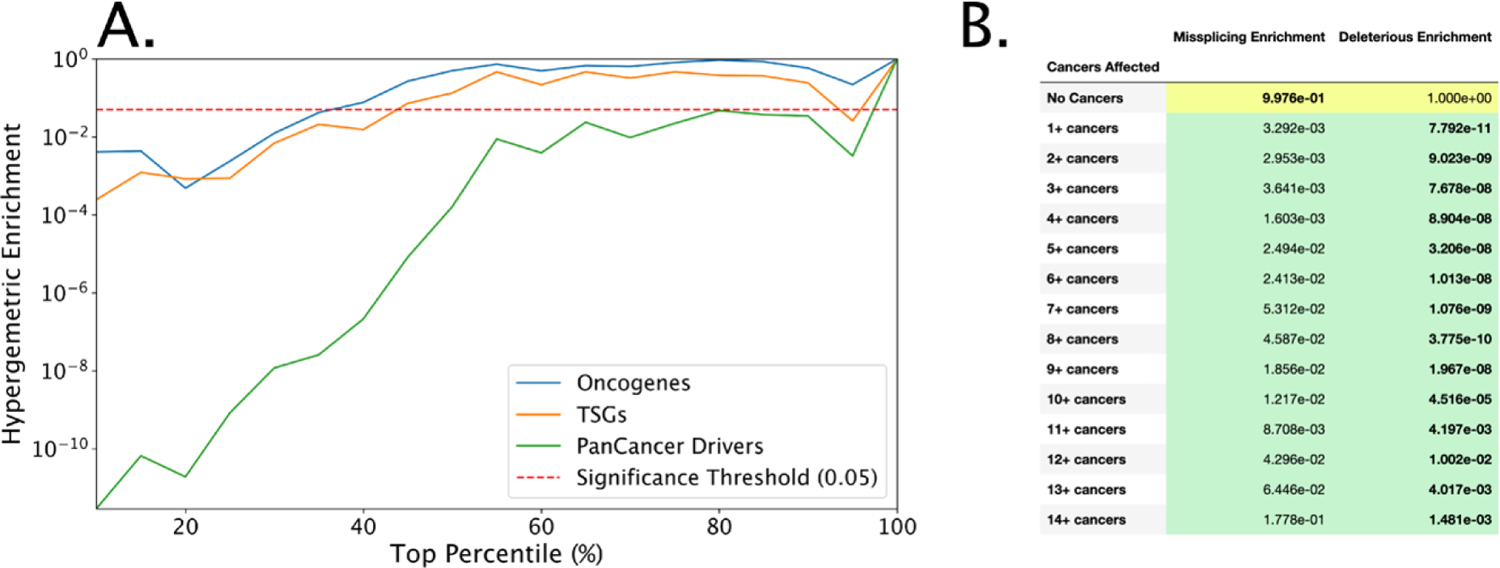
Assessing pan-cancer driver enrichment among deleterious mutations. A. The hypergeometric p-value of the enrichment of known pan-cancer, TSG, and oncogene drivers across the top ranks of overrepresented genes shows that pan-cancer genes are better captured by *Oncosplice* scores. B. The hypergeometric p-value of the enrichment of known pan-cancer across genes that are overrepresented in missplicing and deleterious missplicing mutations across varying numbers of cancer types.

Future cancer treatments and research will be directed toward genes with strong evidence of a potential role in pathogenic mechanisms. Since it has been shown that *Oncosplice* can capture the enrichment of mutations within canonical cancer drivers and TSGs, we can also use this approach to suggest novel cancer genes by looking at those with the highest enrichment of deleterious missplicing events. Therefore, we propose a novel set of potential cancer drivers. This list includes 490 terms included in the top 5% of overrepresented genes among deleterious missplicing mutations. Out of these proposed genes, 49 are canonical pan-cancer drivers. **Figure 6** describes the enrichment of the proposed genes. In essence, these genes can be considered vulnerable to damaging forms of missplicing events and have a role in cancer mechanisms. As seen in **Figure 7A**, the proposed cancer drivers come from the same distribution of all genes in terms of the number of mutations they contain, ensuring selection was not dependent on trivial factors. Many relevant cancer-related molecular functions defined by gene ontology gene sets are strongly enriched within this gene set, including GTPase activity (adjusted hypergeometric p-value: 6.6E-13), G-protein activity (adjusted hypergeometric p-value: 7.4E-6), and helicase activity (adjusted hypergeometric p-value: 1.9E-3)^59^. An unabridged compilation of GO-enriched terms is available in the **Supplementary Table 5**.

**Figure 6.**
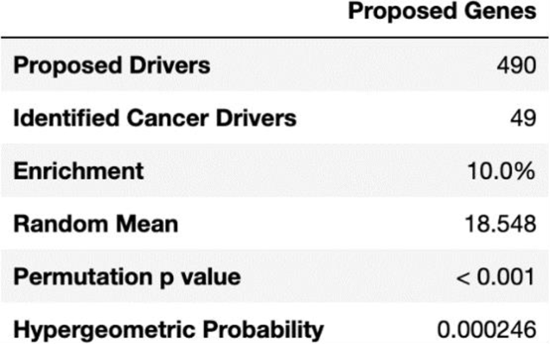
Characteristics of proposed cancer drivers. The list of proposed cancer-related drivers is enriched for known cancer genes.

**Figure 7.**
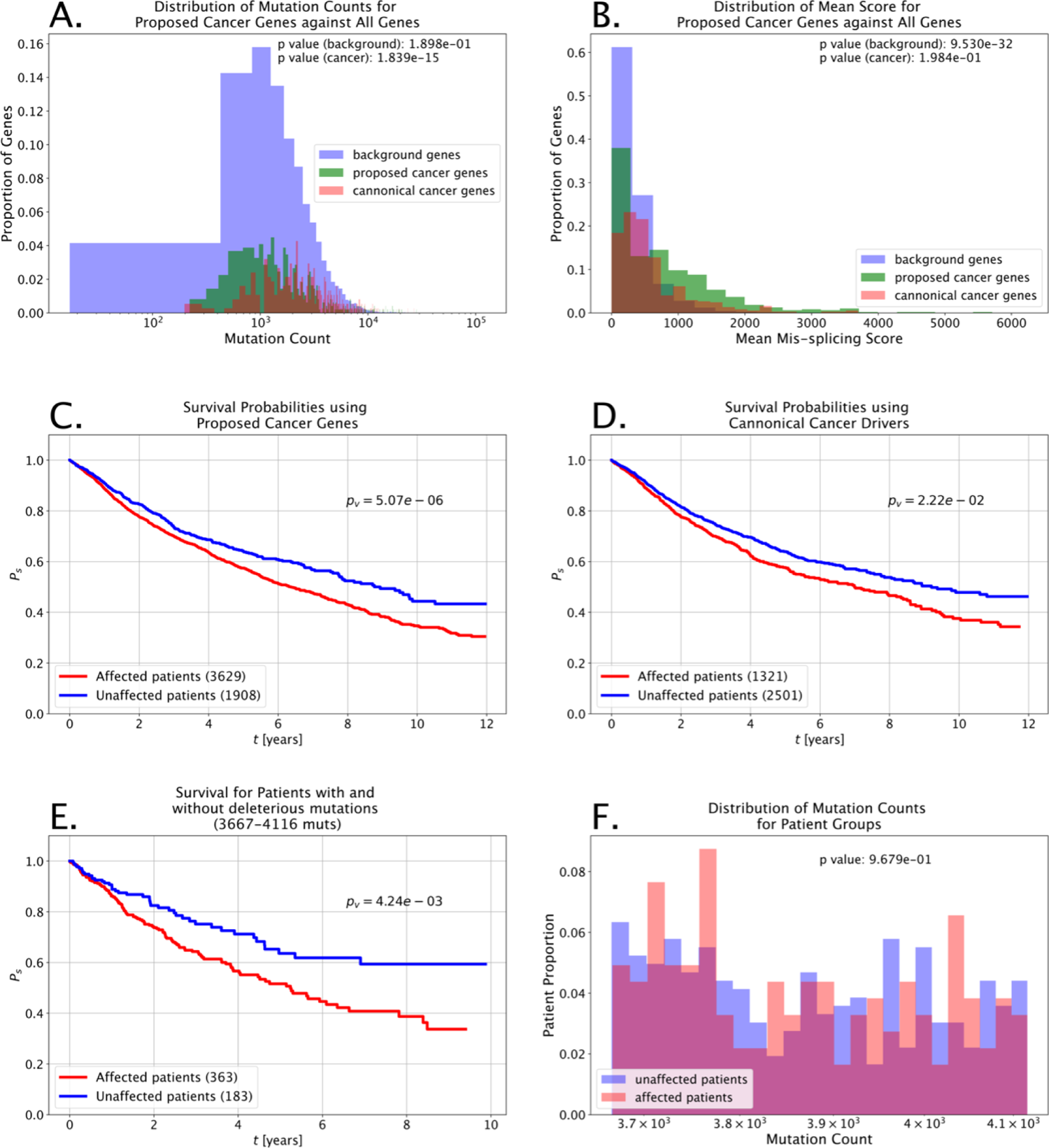
Clinical utility of deleterious missplicing mutations. A. The distributions of mutations per gene for the sets of all genes analyzed, canonical cancer drivers, and the proposed cancer genes show that the proposed genes come from the same distribution as the background gene set rather than having been selected based on trivial characteristics such as mutation volume. B. While the mutation volume for the proposed cancer drivers is not significantly different from all genes analyzed, the pathogenicity of the mutations found in these genes is significantly higher. C. Kaplan Meier survival probabilities for groups of patients defined using mutations within proposed cancer genes. D. Kaplan Meier survival probabilities for groups of patients defined using mutations within canonical cancer genes. E. Kaplan Meier survival probabilities for two groups of patients with similar mutation volumes segmented based on having or not having deleterious mutations. F. Distribution of mutation volumes for patients in groups identified in E shows that the patients do not have significantly different numbers of mutations.

To understand the immediate clinical utility of *Oncosplice* predictions and the proposed cancer drivers, we analyzed survival estimates by identifying patients with deleterious mutations across any of 375 known cancer genes against patients without missplicing mutations in those same cancer genes. We ran similar trials where the known cancer genes were replaced with equally sized sets of genes pulled from the novel 490 proposed genes. As can be seen in **Figure 7C-D**, the segmentation of Kaplan Meier survival estimates for patients using the modified gene list is significantly stronger. This indicates that the novel genes may provide immediate clinical prognostic value. Moreover, we conducted trials to control for the mutation volume across patients by segmenting cases into two groups: those with at least one gene affected by a deleterious mutation and those with no genes affected by deleterious mutations. We then compare the survival probabilities for groups of patients such that there is no significant difference between the mutation volume distributions for the affected and unaffected patients in the subset. In many instances, there was no meaningful difference in survival, though when a significant difference was observed, it was the patients afflicted by deleterious mutations that had more pessimistic outcomes. **Figure 7E** shows the survival probabilities for 546 patients with between 3,667 and 4,116 total mutations. Patients with deleterious mutations have significantly worse survival odds than those without. Moreover, **Figure 7F** shows that the patient groups do not have significantly different mutation volumes and that the segmentation is not reliant on trivial factors. In general, data related to survival is troublesome to work with due to missing values and worsening longitudinal record consistency. Regardless, these results indicate that *Oncosplice* identifies mutations with relation to patient outcomes. Several other analyses in which deleterious missplicing mutations indicate poor survival are available in **Supplementary** Figure 1.

## DISCUSSION

In this work, we have evaluated the signals captured by *Oncosplice*, a novel pipeline with a highlighted utility in characterizing cancerous apparently silent missplicing mutations. We have found that the model identifies many promising patterns in the underlying data across millions of mutations, not the least of which is competitive pathogenicity prediction based solely on estimated aberrant proteomes. Pathogenicity prediction is well-established, and most methods offer high prediction performance, yet the methodology now becomes increasingly important as we try to computationally understand the mechanisms through which variants are deleterious; *Oncosplice* is the only model considered that offers so much in this regard. We have also shown that while *SpliceAI* is an important submodule of the tool, the novelties of our approach add significant predictive power. While *Oncosplice* offers a simple way of identifying mutations based on a measurement of the difference between healthy and aberrant proteomes, we make minimal assumptions about these scores’ precise biological significance. Despite the unclear downstream impact of these grades, there is strong evidence suggesting a large portion of missplicing events result in early termination codons and truncated proteins due to frameshifts introduced by intron retention, exon skipping, or alternative splice site usage; extreme cases of this profile are clearly involved in oncogenesis or disease progression.

When studying cancer at the genomic level, we must remember that data is generally noisy and biased. Annotations to reference transcripts and proteins are hardly conclusive and are frequently updated. New splice sites are also regularly discovered. The inherent tendency of WES to cover CDSs while limiting the availability of untranslated regions or deep intronic sequences also means we are missing many mutations^60^. 98% of the genome is technically invisible through WES, though practically, this is complicated by unpredictable off-target effects. Ultimately, the more true mutations available across all gene regions, the better we can characterize possible splicing aberrations. To improve the analysis enabled by *Oncosplice*, an emphasis must be placed on sequencing non-exonic regions, and increased attention must be given to genes beyond those marked as cancer drivers, which typically have disproportionately larger quantities of verified mutations due to the knowledge of their importance. Still, the statistical power provided by the massive number of mutations analyzed allows generalized insight into aberrant splicing in cancer. Moreover, the splicing-modified gene annotations *Oncosplice* provides allow for rapid analysis of focused case studies.

*Oncosplice’s* ability to predict and reconstruct splicing events that are observed in RNAseq is valuable since this data is expensive and time-consuming to develop. It is hardly trivial to estimate transcriptomes computationally and while there is much room for improvement in terms of predicting true transcripts, those *Oncosplice* generates clearly have biological significance.

Further comparison between predicted aberrant transcripts and those observed in RNAseq will allow for fine-tuning of the model and may lead to isoform likelihood estimates that describe the actual usage of noncanonical splice sites more accurately.

In evaluating this proof of concept, we were also forced to control the testing environment and focus the scope of the investigation on mutations in isolation. Considering the relative success, one highly relevant future direction is to model genes with all simultaneously observed mutations together, as their cumulative effects may change splicing patterns further. This is a task no splicing-related pathogenicity predictor is capable of. Additionally, this tool is not limited to cancer and can be applied to any disease or phenotype cohort, including those representing rare or neurodegenerative diseases. Since *Oncosplice* is unrestricted to any genomic region, we can also explore mutations that no other splicing-related pathogenicity predictor is yet able to address – UTR mutations and nonsilent variants with dual effects. Ultimately, the proposed pipeline serves as a convincing proof of concept showcasing the utility that modeling splicing (and other gene expression mechanisms in general) can have in studying a new dimension of cancer. Splicing is only one of several critical gene expression processes highly impacted by known and unknown apparently silent mutations. Expanding our understanding of how cancer initiates and progresses will require a pivot in attention toward this class of variants that have thus far eluded aggressive research.

## MATERIALS & METHODS

### Identifying missplicing mutations with *SpliceAI*

First, per-nucleotide splicing probabilities are predicted using *SpliceAI*, a deep residual neural network that confidently predicts splice site probabilities for each residue in a sequence based on 10,000 nucleotides of flanking context^26^. The model is capable of splice-site identification with 95% top-k accuracy on arbitrary pre-mRNAs^26^. *SpliceAI* is used within *Oncosplice* to identify changes to splice site usage. Whether a mutation causes aberrant splicing can be estimated using *SpliceAI* in tandem with reference genome annotations by tracking the changes in *SpliceAI* probabilities that nucleotides and splicing junctions near a mutation experience. The four primary events *Oncosplice* detects are missed and discovered acceptors and donor sites. For a mutation of interest, if the donor or acceptor probability of a nearby site decreases by 0.5 or more and that nucleotide is an annotated splice site*, Oncosplice* interprets a missed splice site. If the donor or acceptor probability of a site increases by more than 0.5 and the nucleotide is not an annotated splice site, *Oncosplice* interprets a cryptic or discovered splice site. While it is possible for *SpliceAI* to detect splice sites that have not been formally annotated, they are not considered as there would be no way to assess the quality of these predictions. Higher-order events – including skipped exons and retained intron – are inferred from predicted transcripts.

We look for changes in splicing within a segment 2,500 nucleotides upstream to 2,500 nucleotides downstream of each mutation site. Each mutation is analyzed in isolation, regardless of other mutations that may be concurrently observed in a gene for a given patient. We use 0.5 as a threshold for missplicing detection, a parameter that was validated based on RNAseq data and is the recommended *SpliceAI* parameter^26^. We justify using *SpliceAI* over alternatives such as *MaxEntScan* in the **Supplementary Discussion**.

### Modeling Variant Transcriptomes and Proteomes

Each mutated gene considered by *Oncosplice* has reference genome annotations describing the blueprints for constructing its mature mRNA transcripts and proteins. This data is freely accessible from the *Ensembl* annotation database. Because *SpliceAI* does not consider the schema of all transcripts and donor-acceptor configurations that are biologically observed in each gene, it is not obvious how splicing events can be incorporated into transcripts. Consider, for instance, the simple case of two adjacent cryptic donors.

We use a greedy algorithm that operates on minimal assumptions to handle these situations. This method takes as input a pool of splice sites – reference and predicted alike – that reside within a pre-mRNA transcript’s boundaries. The algorithm follows four rules:

1. Introduce and connect adjected nodes sequentially from 5’ to 3’.
2. Splice sites of the same type cannot be connected.
3. Adjacent splice sites of the same type are equal but exclusive options for connection continuation.
4. Generated splice paths must start with a donor and end with an acceptor.

These guidelines provide an effective construction strategy that is not dependent on unavailable experimental knowledge. The algorithm is not forced to create a single speculative isoform but can generate multiple possible mRNA transcript options, as illustrated in **Figure 1A**. In fact, due to the dynamic and stochastic nature of splice site usage, multiple variant transcripts may be produced, albeit at varying levels. This algorithm handles splice sites at the transcript level and does not require information regarding mutually exclusive exons, cassette exons, or documented alternative boundary usage. Once a mature mRNA transcript is defined, translation is modeled computationally. We consider additional influences from mRNA decay mechanisms^61,62^ and alternative translation initiation site (TIS) usage^63^. Depending on the placement of a discovered site, the span of the transcript may be increased several times over, creating a very long, nonsensical exon. The biological likelihood of such an event occurring is quite low, and even in the case that it was generated by the splicing process, there would likely be some decay mechanisms that would suppress the lifespan of such abnormal transcripts. Transcript isoforms with novel exons longer than 2,000 nucleotides are discarded to account for this. This threshold was selected based on the knowledge that less than 1% of reference human-observed exons exceed 2,000 nucleotides in length.

After obtaining variant mature transcripts, the last major gene expression step is translation. Each transcript in the dataset contains one canonical translation initiation site (TIS) and one canonical translation termination site (TTS). Translating predicted mRNAs may seem trivial. However, untranslated region (UTR) boundaries available in reference transcript annotations may not be usable in variant transcripts. If a reference TIS is disturbed, then a new site is predicted using *TITER*^63^, a deep learning model that predicts optimal TISs based on sequence context, as well as Kozak context score and RNA folding energy. In the case that the reference termination codon is interrupted, or an upstream frameshift renders it unusable, a new TTS is defined by finding the first in-frame canonical termination codon.

### Quantifying the functional divergence of aberrant proteins

Global pairwise alignment provides a good proxy for measuring the similarity between a healthy and predicted variant protein, such as those predicted by *Oncosplice*. In the context of this investigation, a proper alignment must be selected carefully. In aberrant splicing, blocks of nucleotides are apparently inserted or deleted. We consider this by increasing the cost of opening gaps in the pairwise alignment while minimizing the cost of extending gaps. *Biopython’s* pairwise alignment functions are used; we assign one point to aligned amino acids, deduct one point from mismatched amino acids, deduct three points upon gap opening, and deduct no points when extending gaps^64^. Moreover, gaps aligned to the end of the reference protein are not penalized so that truncating events are more visible to the algorithm. These parameters prevent *ad-hoc* alignments with multiple illogical gaps and mismatches that serve only to maximize the alignment score.

While effective, pairwise alignment is naïve since different amino acids in a protein are of varying importance. Certain residues play crucial roles in protein structure or function, and others are involved in neither. One way to ascertain the important domains in a protein is via evolutionary conservation. Such an approach uses the entropy observed for each amino acid residue in aligned homologous proteins to estimate variability and infer functionality. We use *Rate4Site* – a probabilistic evolutionary conservation score calculator that uses Bayesian estimation to obtain relative mutation rates for each position in a multiple sequence alignment (MSA) of homologous proteins based on phylogenic trees^65^. We used *Rate4Site* to process amino acid MSA files for 100 organisms aligned to reference human proteins obtained from UCSC’s database^66^, resulting in a database of per-amino acid evolutionary rates for 109,951 human proteins.

As seen in **Figure 1C**, using pairwise alignment, we can determine the exact positions that are deleted, inserted, and mismatched between the reference and variant protein. Using conservation scores, we can more accurately weigh each position’s importance in the reference sequence. In calculating the magnitude of the functional divergence between two proteins, we consider *W* as a typical protein domain length. This value was obtained by taking the median of all functional domains across available proteins accessible through *InterPro*^67^ – 75 amino acids. *DW* is defined as the length of a detected deletion and *IW* is defined as the length of a detected insertion. *C*(*i, W*) is the mean conservation score of a window of length *W* surrounding a position *i* in the protein. *C*^∗^(*W*) denotes the maximal mean conservation score of a window of length *W* in the analyzed protein.

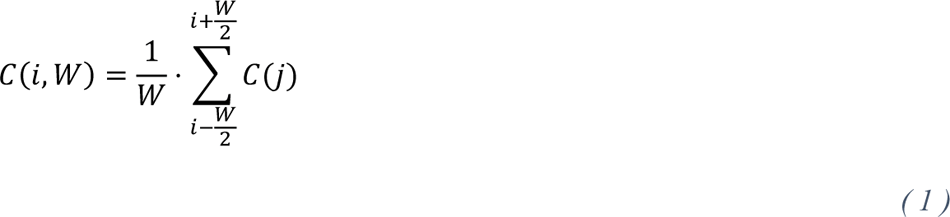

We let *c*(*i, W*) denote 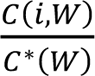, the normalized and smoothed conservation vector.

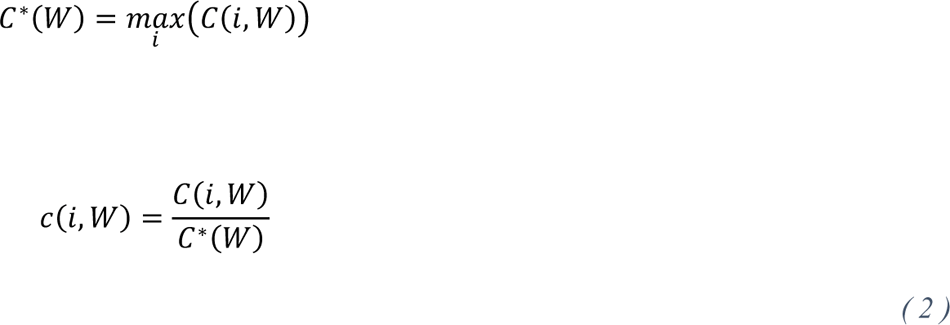

Next, we calculate the value of the deletion-derived functional loss for the deletion of *IW* at position *i* as:

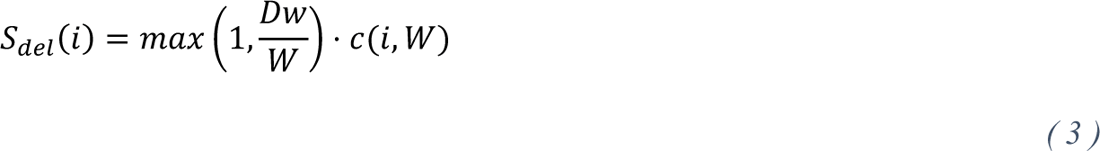

Then we obtain the insertion-derived functional change for the deletion of *iW* at position *i* as:

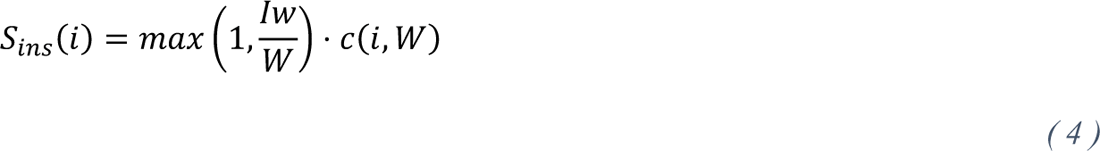

The total penalty for all the deletions and insertions observed in a particular protein is computed using a sliding window of size *W* conflating across deletion and insertion penalties as follows:

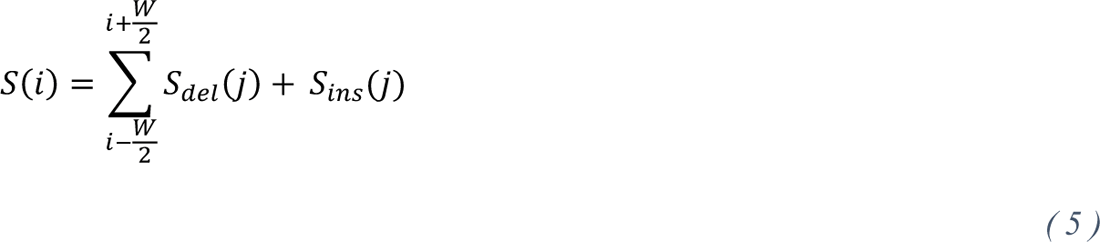

The final score for the respective protein comparison is taken as the maximum value of the penalty vector.

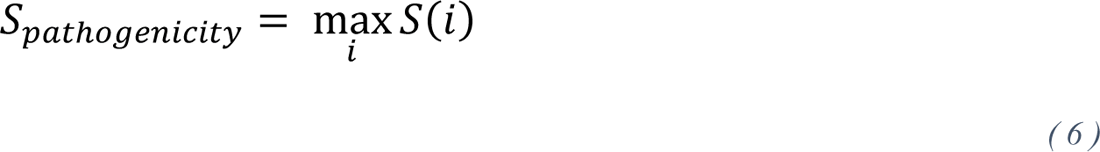

This algorithm is applied to each transcript isoform for a mutated gene. To aggregate these scores into one concise variant descriptor, we implement a weakest-link strategy that obtains the average score for each transcript of a mutated gene across all its predicted isoforms and then assigns the highest score across those transcripts to the mutation. This strategy, shown in **Figure 1D**, describes a mutation by the most dysfunctional protein it generates.

### Benchmarking *Oncosplice* using allele frequencies

To quantify the significance of the overlap between the missplicing mutation dataset and the null dataset, we first find the overlap, or the number of mutations in the missplicing subset that also occur in the null mutation set: *N*^null^_missplicing_. The total number of true missplicing mutations in our variant dataset is denoted as *N*_missplicing_. The pool of all unique mutations observed in the full variant dataset is *S*_unique_. For permutation testing, we perform 1,000 iterations of the following procedure:

1. Create a randomized subset of mutations by selecting *N*_missplicing_ mutations at random from *S*_unique_. This is our fake, randomized subset of missplicing mutations.
2. For iteration *i, N*^fake^_missplicing_ (*i*) is the quantity of mutations in the randomized missplicing mutation set that also occur in the null dataset.

The number of missplicing mutations we expect to occur in the null dataset by chance is the mean of all *N*^fake^_missplicing_ values. The *p value* of the true *N*_missplicing_^null^, divided by the number of conducted iterations.

The hypergeometric probability of obtaining an equal or smaller overlap in null observed mutations within the missplicing subset is computed using the following equation:

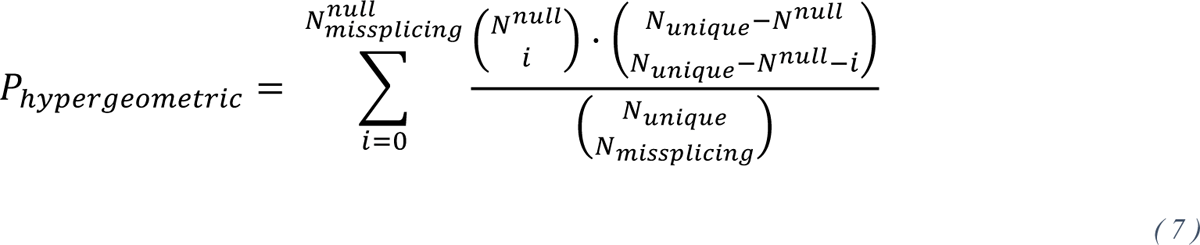

*N*^null^_missplicing_ = number of mutations that are mis-splicing and in null set

*N*^null^ = number of null occurring mutations

*N*_unique_ = number of unique mutations in whole dataset

*N*_missplicing_ = number of mis-splicing mutation

We perform similar permutation and hypergeometric tests when gauging the significance of null depletion in the deleterious missplicing subset, only differing in the set from which we sample our random mutations (we test the depletion relative to the missplicing subset in order to isolate the novel components without *SpliceAI*). Similar procedures are conducted several times across this investigation.

### Benchmarking *Oncosplice* using clinical pathogenicity

*ClinVar* data are parsed and binned into a set containing variant-identifying features (chromosome, mutation position, reference allele, and variant allele) along with their clinical significance and associated disease ontology terms. Clinical significance terms can take on several values though we retain only those with the following tags: “pathogenic”, “likely pathogenic”, “pathogenic/likely pathogenic”, “benign”, “likely benign”, “benign/likely benign”, “uncertain significance”, and “conflicting interpretations”. For simplicity, all values are grouped into “pathogenic” (terms 1-3), “benign” (terms 4-6), or “ambiguous” (terms 7-8) categories.

A joining operation is conducted between our unique cancer mutations and the *ClinVar* data on the variant-identifying features. We now have available three distinct *ClinVar* associated variant sets: unique mutations, missplicing mutations, and deleterious missplicing mutations. For each subset we determine the number of benign, ambiguous, and pathogenic variants. We also calculate the ratio of pathogenic to benign mutations. We measure the success of each subset by the magnitude of this metric.

The significance associated with the pathogenic-to-benign ratio in the missplicing subset is defined by permutation testing; we randomize equally sized subsets of variants by sampling from all unique *ClinVar*-overlapping mutations and check how many randomizations result in a pathogenic-to-benign ratio that is equal or greater. The statistical significance associated with the deleterious missplicing subset is calculated similarly by sampling from the missplicing subset in order to isolate the power of *Onco-splice* novelties from *SpliceAI’s* predictive power.

### Comparing *Oncosplice* to other pathogenicity predictors

We compare the performance of *Onco-splice* against seven alternative pathogenicity predictors, six of which are splicing-specific. To this end, we obtained pre-computed sets of mutations for *CADD, S-CAP, TraP,* and *IntSplice2. MMSplice, RegSNPs-Intron, and RegSNPs-Splicing* did not have sets of pre-computed mutations available, so inference was performed on relevant subsets of the *ClinVar* dataset. The ROC for each tool was obtained using Python’s *sklearn* library. The positive predictive value (PPV) for sets of mutations was obtained by taking all the true pathogenic variants among deleterious classifications and dividing that value by the size of the set of deleterious classifications. Correlations between any two tools were obtained by taking the subset of intersecting variants between those tools and finding the Pearson correlation between the scores of those variants. For tools that grade orthogonal variants, we see that there is no correlation value. For example, *RegSNPs-Intron* and *RegSNPs-Splicing* cannot grade the same variants; hence, no correlation is obtained.

### Measuring driver gene enrichment

To first obtain a baseline estimate as to whether cancer genes contain higher ratios of deleterious mutations compared to other genes, we calculate the significance of the average ratio of deleterious mutations to unique mutations across cancer genes and compared that value to non-cancer gene ratios.

We employ permutation testing by performing the following procedure 10,000 times:

1. For each gene *g*, calculate the rate of deleterious mutations as *R*^del^_g_ = N^del^_g_/N^tot^_g_ where *N*^del^_g_ is the number of deleterious mutations in *g* and *N*^tot^ is the number of total mutations in *g*.
2. Obtain the mean of all *R*^del^_g_ for known cancer genes and call this *R*^del^_cancer_
3. Randomize a group of genes of size *N*_cancer_ where *N*_cancer_ is the number of known cancer genes used in Step 2.
4. Obtain the mean of the randomized gene group’s *R*^del^_g_ in iteration *i*, called *R*^del^_random_ (*i*).

After performing these steps, determine how often these randomizations result in *R*^del^_random_ that is greater than or equal to *R*^del^_cancer_ by calculating:

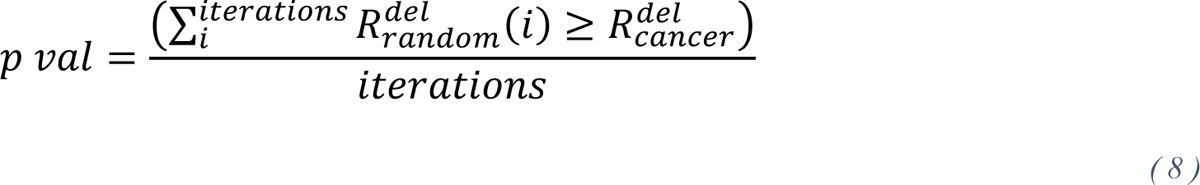

Our objective is to validate *Onco-splice’s* ability to identify cancer-driving mutations by showing that genes disproportionately overrepresented among deleterious missplicing mutations are enriched with known cancer genes. Yet, known cancer genes have more mutations than non-cancer genes and this bias must be addressed. Therefore, to find genes that are overrepresented by deleterious mutations while mitigating mutation volume bias, we design the following procedure which operates on any arbitrary pool of mutations.

1. We determine the number of unique mutations for each gene – *N*_unique_. Based on this count, we divide genes into 5 quantile groups having similar mutation volumes.
2. For each gene, we determine the count of missplicing (*N*_mis_) and deleterious missplicing (*N*_del_) mutations and further develop these values into missplicing and deleterious missplicing mutation ratios as:

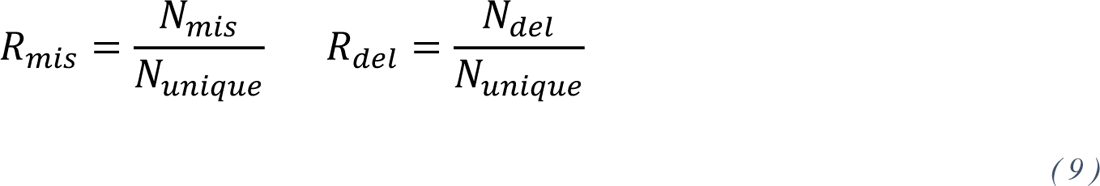
3. Within each quantile group, we sort genes based on one of the target ratios. To study, say, the top 5% of all overrepresented genes in the deleterious subset (as is done to identify the proposed set of novel cancer drivers), we select the top 5% of genes from each quantile based on *R*_del_.

Once a set of overrepresented genes is obtained, the level of cancer gene enrichment can be obtained using permutation and hypergeometric testing as described previously. We follow a similar strategy when finding cancer-specific enrichment by performing this procedure on the sets of mutations found in each cancer type. We track the genes that are overrepresented in cancer type and then count the total projects that each gene is found to be overrepresented in.

### Estimating patient survival

To show the clinical value of the proposed cancer genes and *Onco-splice* we generate two sets of patients: one defined as the affected case set and one as the unaffected case set. In one survival analysis, the affected case set is determined by finding all the patients in the cohort who have one deleterious mutation in a defined set of cancer genes. The unaffected case set is determined by finding all the patients in the cohort who have no missplicing mutations in the same defined set of cancer genes. The set of cancer genes in the control experiment is defined as 375 known pan-cancer genes. The set of cancer genes in the variable experiment is defined as a random set of 375 genes from the proposed cancer gene set (we sample 375 genes randomly to ensure that there is no bias related to the size of the gene set. For each experiment (or set of affected and unaffected patients), we calculate the survival rates and the significance of their differences for 10- or 12-year survival using Kaplan Meier survival estimation. This analysis is robust to changes in the size of the gene set and the length of survival time. The significance of the test set is always stronger than the control set, regardless of the subset of 375 proposed cancer genes selected.

In a second survival analysis, we aim to validate identified deleterious mutations while controlling for bias related to mutation volume in the selection of patients for each group. To this end, we generate two sets of patients: those who contain at least one gene affected by a deleterious mutation and those who are not affected by a deleterious mutation. These two sets of groups have a very strong difference in the distribution of mutation volumes, with the affected patients containing many more mutations than the unaffected case group. To understand if the signal persists when eliminating the mutation volume bias, we look at subsets of patients that contain no significant difference in their distributions of mutation volumes by binning based on percentiles.

### Identifying Gene Ontology (GO) terms

Gene enrichment analysis was performed using *g:Profiler*^59^, a web tool that performs hypergeometric enrichment analysis for a target gene set against a background gene set using a database of GO terms and their associated sets of terms. The primary list of genes was defined as the set of proposed novel cancer drivers. The background set is defined as all the genes with mutations that were studied. After running the analysis, *g:Profiler* provides adjusted p values for each identified term. This tool is updated with the latest GO terms and sets.

## DECLARATIONS

### Data Availability

The data used in this study was generated by The Cancer Genome Atlas (https://www.cancer.gov/tcga) and can be downloaded from the genomic data commons (https://portal.gdc.cancer.gov/).

MSAs were accessed from UCSC (https://hgdownload.soe.ucsc.edu/downloads.html#human).

Genome annotations were accessed from *Ensembl* (https://www.ensembl.org/info/data/ftp).

## Acknowledgments

Not applicable

## Author Contributions

TT conceived this study. NL analyzed the data. TT supervised the study. NL and TT wrote the paper. All authors read and approved the final manuscript.

## References

1. Weinstein, J. N. et al. The Cancer Genome Atlas Pan-Cancer analysis project. Nat Genet 45, 1113–1120 (2013).

2. Tate, J. G. et al. COSMIC: the Catalogue Of Somatic Mutations In Cancer. Nucleic Acids Res 47, D941–d947 (2019).

3. Clarke, L. et al. The international Genome sample resource (IGSR): A worldwide collection of genome variation incorporating the 1000 Genomes Project data. Nucleic Acids Res 45, D854–D859 (2017).

4. Stephens, Z. D. et al. Big Data: Astronomical or Genomical? PLOS Biology 13, e1002195 (2015).

5. Cuykendall, T. N., Rubin, M. A. & Khurana, E. Non-coding genetic variation in cancer. Current Opinion in Systems Biology 1, 9–15 (2017).

6. Rheinbay, E. et al. Analyses of non-coding somatic drivers in 2,658 cancer whole genomes. Nature 578, 102–111 (2020).

7. Corona, R. I. et al. Non-coding somatic mutations converge on the PAX8 pathway in ovarian cancer. Nat Commun 11, 2020 (2020).

8. Zhou, S. et al. Noncoding mutations target cis-regulatory elements of the FOXA1 plexus in prostate cancer. Nat Commun 11, 441 (2020).

9. Zhang, X. & Meyerson, M. Illuminating the noncoding genome in cancer. Nature Cancer 1, 864–872 (2020).

10. Waldman, Y. Y., Tuller, T., Sharan, R. & Ruppin, E. TP53 cancerous mutations exhibit selection for translation efficiency. Cancer Res 69, 8807–13 (2009).

11. Gutman, T., Goren, G., Efroni, O. & Tuller, T. Estimating the predictive power of silent mutations on cancer classification and prognosis. *npj Genom*. Med. 6, 1–15 (2021).

12. Diederichs, S. et al. The dark matter of the cancer genome: aberrations in regulatory elements, untranslated regions, splice sites, non-coding RNA and synonymous mutations. EMBO Mol Med 8, 442–457 (2016).

13. Barash, Y. et al. Deciphering the splicing code. Nature 465, 53–59 (2010).

14. Bergman, S. & Tuller, T. Widespread non-modular overlapping codes in the coding regions. Phys Biol 17, 031002 (2020).

15. Bonnal, S. C., López-Oreja, I. & Valcárcel, J. Roles and mechanisms of alternative splicing in cancer - implications for care. Nat Rev Clin Oncol 17, 457–474 (2020).

16. Cao, S. et al. Discovery of driver non-coding splice-site-creating mutations in cancer. Nature communications 11, 5573 (2020).

17. Cartegni, L., Chew, S. L. & Krainer, A. R. Listening to silence and understanding nonsense: exonic mutations that affect splicing. Nat Rev Genet 3, 285–298 (2002).

18. Hansen, T. V. O. et al. The silent mutation nucleotide 744 G --> A, Lys172Lys, in exon 6 of BRCA2 results in exon skipping. Breast Cancer Res Treat 119, 547–550 (2010).

19. Kahles, A. et al. Comprehensive Analysis of Alternative Splicing Across Tumors from 8,705 Patients. Cancer Cell 34, 211–224.e6 (2018).

20. Park, J. W. & Graveley, B. R. Complex alternative splicing. Adv Exp Med Biol 623, 50–63 (2007).

21. Sakai, A. et al. Aberrant expression of CPSF1 promotes head and neck squamous cell carcinoma via regulating alternative splicing. PLoS One 15, e0233380 (2020).

22. Sciarrillo, R. et al. The role of alternative splicing in cancer: From oncogenesis to drug resistance. Drug Resistance Updates 53, 100728 (2020).

23. Supek, F., Miñana, B., Valcárcel, J., Gabaldón, T. & Lehner, B. Synonymous mutations frequently act as driver mutations in human cancers. Cell 156, 1324–1335 (2014).

24. Wang, G.-S. & Cooper, T. A. Splicing in disease: disruption of the splicing code and the decoding machinery. Nature Reviews Genetics 8, 749–761 (2007).

25. Brinkman, B. M. N. Splice variants as cancer biomarkers. Clin Biochem 37, 584–594 (2004).

26. Jaganathan, K. et al. Predicting Splicing from Primary Sequence with Deep Learning. Cell 176, 535–548.e24 (2019).

27. Lee, H. S. et al. Chemical suppression of an oncogenic splicing variant of AIMP2 induces tumour regression. Biochem J 454, 411–416 (2013).

28. Martinez-Montiel, N., Rosas-Murrieta, N. H., Anaya Ruiz, M., Monjaraz-Guzman, E. & Martinez-Contreras, R. Alternative Splicing as a Target for Cancer Treatment. Int J Mol Sci 19, E545 (2018).

29. Sveen, A., Kilpinen, S., Ruusulehto, A., Lothe, R. A. & Skotheim, R. I. Aberrant RNA splicing in cancer; expression changes and driver mutations of splicing factor genes. Oncogene 35, 2413–2427 (2016).

30. Frankiw, L., Baltimore, D. & Li, G. Alternative mRNA splicing in cancer immunotherapy. Nat Rev Immunol 19, 675–687 (2019).

31. Jung, H. et al. Intron retention is a widespread mechanism of tumor-suppressor inactivation. Nat Genet 47, 1242–1248 (2015).

32. Christofk, H. R. et al. The M2 splice isoform of pyruvate kinase is important for cancer metabolism and tumour growth. Nature 452, 230–233 (2008).

33. Lin, J. et al. Base editing-mediated perturbation of endogenous PKM1/2 splicing facilitates isoform-specific functional analysis in vitro and in vivo. Cell Prolif 54, e13096 (2021).

34. Poulikakos, P. I. et al. RAF inhibitor resistance is mediated by dimerization of aberrantly spliced BRAF(V600E). Nature 480, 387–390 (2011).

35. Rentzsch, P., Schubach, M., Shendure, J. & Kircher, M. CADD-Splice—improving genome-wide variant effect prediction using deep learning-derived splice scores. Genome Medicine 13, 31 (2021).

36. Cheng, J. et al. MMSplice: modular modeling improves the predictions of genetic variant effects on splicing. Genome Biology 20, 48 (2019).

37. Gelfman, S. et al. Annotating pathogenic non-coding variants in genic regions. Nat Commun 8, 236 (2017).

38. Takeda, J., Fukami, S., Tamura, A., Shibata, A. & Ohno, K. IntSplice2: Prediction of the Splicing Effects of Intronic Single-Nucleotide Variants Using LightGBM Modeling. Frontiers in Genetics 12, 1232 (2021).

39. Lin, H. et al. RegSNPs-intron: a computational framework for predicting pathogenic impact of intronic single nucleotide variants. Genome Biology 20, 254 (2019).

40. Zhang, X. et al. regSNPs-splicing: a tool for prioritizing synonymous single-nucleotide substitution. Hum Genet 136, 1279–1289 (2017).

41. Jagadeesh, K. A. et al. S-CAP extends pathogenicity prediction to genetic variants that affect RNA splicing. Nat Genet 51, 755–763 (2019).

42. Jung, H., Lee, K. S. & Choi, J. K. Comprehensive characterisation of intronic mis-splicing mutations in human cancers. Oncogene 40, 1347–1361 (2021).

43. Jayasinghe, R. G. et al. Systematic Analysis of Splice-Site-Creating Mutations in Cancer. Cell Reports 23, 270–281.e3 (2018).

44. Nathany, S. & Batra, U. MET: A narrative review of exon 14 skipping mutation in non-small-cell lung carcinoma. *Cancer Research*, Statistics, and Treatment 5, 284 (2022).

45. Chen, S. et al. A genome-wide mutational constraint map quantified from variation in 76,156 human genomes. 2022.03.20.485034 Preprint at 10.1101/2022.03.20.485034 (2022).

46. Jung, H., Bleazard, T., Lee, J. & Hong, D. Systematic investigation of cancer-associated somatic point mutations in SNP databases. Nat Biotechnol 31, 787–789 (2013).

47. Landrum, M. J. et al. ClinVar: public archive of relationships among sequence variation and human phenotype. Nucleic Acids Res 42, D980–985 (2014).

48. Landrum, M. J. et al. ClinVar: public archive of interpretations of clinically relevant variants. Nucleic Acids Res 44, D862–868 (2016).

49. Landrum, M. J. et al. ClinVar: improving access to variant interpretations and supporting evidence. Nucleic Acids Res 46, D1062–D1067 (2018).

50. Dietlein, F. et al. Identification of cancer driver genes based on nucleotide context. Nat Genet 52, 208–218 (2020).

51. Asthana, S., Roytberg, M., Stamatoyannopoulos, J. & Sunyaev, S. Analysis of Sequence Conservation at Nucleotide Resolution. PLOS Computational Biology 3, e254 (2007).

52. Liu, Y., Sun, J. & Zhao, M. ONGene: A literature-based database for human oncogenes. J Genet Genomics 44, 119–121 (2017).

53. Martínez-Jiménez, F. et al. A compendium of mutational cancer driver genes. Nat Rev Cancer 20, 555–572 (2020).

54. Sondka, Z. et al. The COSMIC Cancer Gene Census: describing genetic dysfunction across all human cancers. Nat Rev Cancer 18, 696–705 (2018).

55. Zhao, M., Sun, J. & Zhao, Z. TSGene: a web resource for tumor suppressor genes. Nucleic Acids Res 41, D970–6 (2013).

56. Lawrence, M. S. et al. Discovery and saturation analysis of cancer genes across 21 tumour types. Nature 505, 495–501 (2014).

57. Bailey, M. H. et al. Comprehensive Characterization of Cancer Driver Genes and Mutations. Cell 173, 371–385.e18 (2018).

58. Repana, D. et al. The Network of Cancer Genes (NCG): a comprehensive catalogue of known and candidate cancer genes from cancer sequencing screens. Genome Biology 20, 1 (2019).

59. Raudvere, U. et al. g:Profiler: a web server for functional enrichment analysis and conversions of gene lists (2019 update). Nucleic Acids Research 47, W191–W198 (2019).

60. Barbitoff, Y. A. et al. Systematic dissection of biases in whole-exome and whole-genome sequencing reveals major determinants of coding sequence coverage. Sci Rep 10, 2057 (2020).

61. Singh, P., Saha, U., Paira, S. & Das, B. Nuclear mRNA Surveillance Mechanisms: Function and Links to Human Disease. J Mol Biol 430, 1993–2013 (2018).

62. Garneau, N. L., Wilusz, J. & Wilusz, C. J. The highways and byways of mRNA decay. Nat Rev Mol Cell Biol 8, 113–126 (2007).

63. Zhang, S., Hu, H., Jiang, T., Zhang, L. & Zeng, J. TITER: predicting translation initiation sites by deep learning. Bioinformatics 33, i234–i242 (2017).

64. Cock, P. J. A. et al. Biopython: freely available Python tools for computational molecular biology and bioinformatics. Bioinformatics 25, 1422–1423 (2009).

65. Pupko, T., Bell, R. E., Mayrose, I., Glaser, F. & Ben-Tal, N. Rate4Site: an algorithmic tool for the identification of functional regions in proteins by surface mapping of evolutionary determinants within their homologues. Bioinformatics 18 **Suppl 1**, S71–77 (2002).

66. Nassar, L. R. et al. The UCSC Genome Browser database: 2023 update. Nucleic Acids Res 51, D1188–D1195 (2023).

67. Blum, M. et al. The InterPro protein families and domains database: 20 years on. Nucleic Acids Research 49, D344–D354 (2021).

